# “Puberty age gap”: New method of assessing pubertal timing and its association with mental health problems

**DOI:** 10.1101/2022.05.13.22275069

**Authors:** Niousha Dehestani, Nandita Vijayakumar, Gareth Ball, Mansour L. Sina, Sarah Whittle, Timothy J. Silk

## Abstract

Puberty is linked to mental health problems during adolescence, and in particular, the timing of puberty is thought to be an important risk factor. This study developed a new measure of pubertal timing using multiple pubertal features and nonlinear associations with age and investigated its association with mental health problems. Using the Adolescent Brain Cognitive Development (ABCD) cohort, we implemented three models of pubertal timing by predicting chronological age from i) observed physical development, ii) hormones (testosterone and dehydroepiandrosterone [DHEA]), and iii) a combination of the two, using a supervised machine learning method (n ∼9,900). Accuracy of the new models, and their associations with mental health problems were evaluated. The new pubertal timing measure performed better in capturing age variance compared to a commonly used method, and the physical measure accounted for more variance in mental health, such that earlier pubertal timing was associated with higher symptoms. This study demonstrates the utility for a new model of pubertal timing and suggests that physical maturation may play a predominant role in predicting mental health problems in early adolescence.

## Introduction

Adolescence is the period between childhood and adulthood when individuals acquire the emotional and cognitive skills necessary to gain independence from their parents(1,2). Individuals also progress through puberty, the process of attaining reproductive maturity, during this period. This includes hormonal and physical changes such as body hair growth and gonadal maturation (along with the development of breasts and onset of menstruation in females)(3,4). The progression through puberty has been associated with an increase in susceptibility to a range of internalizing and externalizing mental health problems(5,6), which may reflect the effects of hormones on the central nervous system and/or psychosocial mechanisms related to physical differences from peers(7). Therefore, investigation of individual differences in pubertal processes that consider both hormonal and physical changes may lead to a better understanding of adolescent mental health problems.

While all typically developing individuals progress through the same stages of puberty (based on observable physical changes), the onset and speed of progression can differ across individuals. As such, at any given period during adolescence, there is marked variability in pubertal stage, termed “pubertal timing” (8,9). Importantly, it is pubertal timing – not pubertal stage – that is often linked to the emergence and severity of mental health problems (10), though there are inconsistencies in the literature. A number of studies have found that in females, earlier pubertal timing is associated with internalizing problems (11,12), including depression(13,14), anxiety(15), eating disorders (16), and externalizing behaviors. On the other hand, a number of other studies report null findings regarding internalizing symptoms(14,15). Similarly in males, both early and late timing has been related to internalizing (11 VS 16) and externalizing problems (17 VS 18), while others have failed to identify any associations (9, 15). Small sample sizes and methodological differences between studies may partially explain the inconsistent findings, with variation in the measures used to calculate pubertal timing likely important. Indeed, a meta-analysis of 101 studies found that the method of measuring puberty had a moderating role in the relationship between pubertal timing and mental health problems (4).

Different statistical approaches exist to measure the relative pubertal timing of individuals compared to same-aged peers. A common approach is to regress age from pubertal status (e.g., based on Pubertal Developmental Scale [PDS] scores) in a sample, with residuals reflecting earlier or later maturity relative to the group average (21). However, such approaches only capture individual differences in observable physical development and do not inform us about underlying biological mechanisms that are more reflected in hormone levels (22). Hormone levels provide valuable information regarding the endocrine processes of puberty (23,24). Previous studies have also demonstrated associations between changes in hormone levels and adolescent mental health problems. For example, increasing levels of dehydroepiandrosterone (DHEA) have been linked to internalizing symptoms (25), while testosterone has been associated with externalizing and disruptive behaviors (26). To the best of our knowledge, only one study has calculated pubertal timing with hormone data to predict internalizing behaviors in a sample of 174 females (27). Although this study did not find any significant associations, it is important to investigate this in a larger sample, of both females and males, and to assess associations across different dimensions of mental health problems. Additionally, no previous studies have combined both hormonal and physical measurements when calculating pubertal timing, which may be more sensitive to multiple underlying mechanisms that contribute to mental health problems. Thus, further investigation is needed to develop pubertal timing methods that amalgamate different aspects of puberty in a single model.

In this study, we propose a multivariate method to calculate pubertal timing that draws upon the “brain age” approach (28,29), where an association between multiple neuroimaging variables and chronological age is learned with supervised machine learning methods. Subtracting chronological age from brain age yields a “brain age gap” that reflects brain maturation relative to the group average (26,28). The major benefits of this model include being able to combine multiple features and reducing complex multivariate information to a single parameter. We propose to use a similar model to calculate estimates of pubertal timing by combining multiple puberty-related features. In contrast, alternative strategies either focus on single measures and only capture specific aspects or mechanisms of puberty or calculate the mean of multiple measures but may obscure the relative importance of each item. The second benefit of this method is that it can model nonlinear relationships between multiple measures of puberty and age, which is important given that nonlinear associations between specific features of puberty and chronological age have been observed in previous studies (30,31).

In the current study, we aimed to create a normative model of pubertal timing that combines multiple measures of puberty capturing hormone levels and physical changes. By using a strict cross-validation approach, we ensure that model performance is robust. We compared model performance to a common linear model of pubertal timing (i.e., regressing age from total PDS score). We also compared the combined normative model against two unimodal models employing a similar supervised machine learning approach, one that uses hormonal measures alone and another that uses physical measures only. Finally, we examined the association between each pubertal timing model and multiple dimensions of mental health problems. We hypothesized that the combined normative model of pubertal timing, which uses both hormonal and physical measures, would provide a more accurate prediction of age and better prediction of mental health problems compared to either of the unimodal (hormonal or physical) normative models or the (traditional) linear model of puberty. We additionally hypothesized that early pubertal timing would be associated with increased mental health problems in females and males. Based on a recent meta-analysis, we hypothesized significant effects of similar magnitude for internalizing and externalizing problems, while no significant effects were expected for attention problems (4). Prior findings of sex differences are mixed, and as such we did not propose specific hypotheses here.

## Materials and Methods

To promote reproducible open research practices, all analyses conducted as part of this article are made publicly available in a git repository (“https://github.com/Niousha-Dehestani/Puberty-age”). In addition, more detail about participants, measurements and analysis is provided in the supplementary materials.

### Participants

Participants were drawn from the ongoing, longitudinal, Adolescent Brain Cognitive Development (ABCD) Study (https://abcdstudy.org/). Data was collected from ∼11,500 children at baseline (47% females, age 9-10 years old) from 21 sites across the United States, with annual data collection thereafter (see Supplementary Table 1 for demographic information). Data from baseline and 3 annual follow-up waves were used in the current analyses. We excluded participants who had a mismatch between their biological sex (collected per visit with the salivary sample) and their self-reported gender as well as those with missing values in biological sex (See Supplementary Information (SI) Appendix S1 for details of data cleaning procedures). The exact sample sizes utilized in each analysis are reported in detail below.

### Measures

#### Pubertal Development Scale

The Pubertal Development Scale (PDS) measures observable physical signs of puberty. It includes items on height, body hair, and skin change in both sexes, as well as the onset of menarche and breast development for girls, and facial hair and voice changes for boys. Items are rated on a Likert scale from one to four points (“had not begun” to “already complete”). Onset of menarche was a binary variable (yes/no response) that was converted to one for “no” and four for “yes”. The PDS can either be collected via self- or parent-report. However, each of these measures have their own limitations. The parent-report has good correspondence to clinician ratings, though this correspondence is lower in males (32). Conversely, the self-report is less accurate for individuals who are in the lower or upper pubertal stages as they tend to report toward the mid stages. Therefore, some studies have recommended using the parent-report PDS, especially in late childhood or early adolescence (33), and accordingly, the current study utilized this version. In this study we used individual PDS items rather than the average PDS score typically used in prior literature. This allowed us to capture the unique relationship between each item and our outcomes of interest, consistent with previous literature that has found that separate PDS items exhibit differential relationships with, e.g., brain structure (34).

#### Hormones

DHEA and testosterone (TST) levels were measured via salivary hormone samples assayed by Salimetrics. Although estradiol was also measured, it was not used in the current analyses due to it only being available for females and having excessive missingness (n = 780). The hormone data was cleaned based on the protocol that published recently(35), which involved removing the confounding effects of collection time, duration of collection, wake-up time on collection day, having exercised before collection, and caffeine intake (with a linear mixed effect model, see SI for more details). Further information on the reliability and overall quality of the hormone data can be found in protocol (32).

#### Body Mass Index (BMI)

BMI was calculated as the average of two weight and height measurements per visit, assessed by the researcher. Next, BMI standard deviation scores (BMI z-scores) were calculated relative to age and sex, with reference to the CDC 2000 Growth Charts (36).

#### Sociodemographic Variables

Five categories of race/ethnicity were coded: White, Black, Hispanic, Asian, and Other/Multi-race. Additionally, household income and education were obtained as measures of family socioeconomic status (SES) that were collected in a parent report.

#### Mental health problems

The Child Behavior Checklist (CBCL) (age 6 to 18 form; (37)) was used to measure parent-reported mental health problems. CBCL includes 8 syndrome scales; namely, Anxious/Depressed, Withdrawn/Depressed, Somatic Complaints, Social Problems, Thought Problems, Attention Problems, Aggressive Behavior, and Rule-breaking Behavior as well as three broad summary scales of externalizing, internalizing, and total problems.

### Statistical Analysis

#### Calculating pubertal timing

Inheriting fundamental concepts from the literature on “brain age” (26), “puberty age” was computed using supervised machine learning. The model was trained to learn the relationship between physical and hormonal measurements of puberty (specifically, each PDS item, DHEA levels and TST levels) and chronological age, separately in males and females. Thereafter, the model was used to predict chronological age from pubertal measurements in an independent test sample. The effect of age was subsequently regressed from these predictions to adjust the bias created by regression toward the mean (RTM) (For more details see SI, Appendix S3, Figure S2 and Figure S3, and (38)). The bias-adjusted prediction of an individual’s chronological age from pubertal measurements is termed “puberty age”. Further, the residuals of the prediction model (after subtracting chronological age from puberty age) are referred to as the “puberty age gap”, which we use as a dimensional measure indicative of relative pubertal timing. A positive puberty age gap is interpreted as a sign of earlier pubertal development compared to the normative age and sex-matched group, while a negative gap reflects relatively delayed pubertal development (see, Appendix 1, Figure S1).

We implemented a Generalized Additive Model (GAM) for the prediction of age (response variable) from pubertal measurements, which utilized TST, DHEA, and each of the PDS items as multivariate predictor variables. To remove the potential impacts of familial relations and repeat assessments of each individual, the sample was stratified to randomly keep a single observation for each family (i.e., across waves and siblings). For details on data processing, such as the procedure for dealing with missing values, see SI Appendix S1 and Table S1. Additionally, to ensure the model was trained on a typically developing sample; using the CBCL DSM-oriented scales (consistent with DSM diagnostic categories), we excluded individuals above the threshold (symptom’s score >60) for affective, anxiety, somatic, opposition defiant and conduct problems, as well as ADHD. The remaining participants who had symptom scores <60 were included in the training of models of puberty age (N = 4,949 (2,439 females)). However, a representation of the whole sample was used for testing (N = 9,919 (4,725 females)).

As GAM fits smooth nonlinear curves in the form of spline functions, it is expected to outperform commonly utilized linear methods of pubertal timing measurement, given the nonlinear relations previously reported(30). Model training was performed within a nested 10-fold cross-validation design (for each fold in the outer loop, 90% of samples were used for model training and 10% for testing). Inner loop hyperparameter tuning was performed by a grid search for optimal regularization penalty on each term (i.e, using GAM) to minimize the estimated prediction error in the training sample (generalized cross-validation (GCV) score). To understand the contribution of different indices to the measurement of pubertal timing, we implemented three alternative models to estimate “puberty age”. The first approach only used hormones (DHEA and TST) as features to predict chronological age, the second only used PDS items, and the third combined hormone and PDS items. The predictions of age were named *hormonal* puberty age, *physical* puberty age, and *combined* puberty age, respectively. In all three models, the residuals of age prediction indexed “puberty age gap”, which reflects pubertal timing. We also used the partial dependence function in GAM models that can reflect the importance of each feature in the combined puberty age model (for more detail, see SI, Appendix S4 and Figure S4).

#### Comparison of “puberty age” models

We compared the performance of the three alternate puberty age models using Pearson’s correlation between predicted and chronological age and median absolute error (MAE) averaged over the 10 cross-validation folds. Additionally, we used a non-parametric paired t-test (Wilcoxon signed-rank test) to statistically compare the model performance of different models based on the absolute error of predictions.

#### “Physical puberty age” compared to a traditional pubertal timing model

The current study also compared the out-of-sample performance and accuracy of the physical puberty age model with the most common traditional pubertal timing model: linearly regressing age from average PDS score. In order to draw a comparison between the new model and the traditional approach, the performance of the physical puberty age model was contrasted with the traditional approach. The physical model was selected (rather than the hormonal or combined model) to ensure a fair comparison of two approaches that measure pubertal timing from the same input features (i.e., PDS). In order to conduct this comparison, a linear regression model was used to regress chronological age from the total PDS score (train) and used the fitted model coefficients in unseen data to measure timing (test). Similar to the puberty age model design, a 10-fold cross-validation design was used to measure traditional pubertal timing for the whole sample. This linear model provided an implementation of the traditional model in an out-of-sample prediction paradigm. Model performance was assessed based on the out-of-sample prediction accuracies (quantified by the absolute error of predictions). Similarly, we used the “Wilcoxon signed-rank test” to investigate the statistical differences in model performance.

#### “Puberty age gap” associations with mental health problems

We used linear mixed-effect models (LMM) to investigate associations between each alternate “puberty age gap” measure and different dimensions of mental health problems, in males and females separately. The following formula was tested for each syndrome dimension (i.e., Anxious/Depressed, Withdrawn/Depressed, Somatic Complaints, Social Problems, Thought Problems, Attention Problems, Aggressive Behavior, and Rule-breaking Behavior), as well as three broad scales including total problems, externalizing, and internalizing problems. Sample size for this analysis was N = 9,919 (4,725 (females)) after removing the missing value in CBCL items.

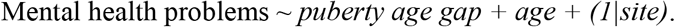

Age was included as a confound (fixed effect) and data collection sites were modelled as random effects. We corrected for multiple tests controlling for False Discovery Rate (FDR) at 5% and reported the FDR corrected p-values in the results. The different “pubertal age” LMMs were compared based on the Akaike Information Criterion (AIC) and a cut-off of 2 was used to indicate evidence for a better model, i.e., the model with an AIC that is at least 2 units smaller is considered a comparatively better model. Furthermore, to investigate whether associations between pubertal timing and mental health problems differed by age, we investigated the interaction effect of age and puberty age gap in predicting mental health problems using the following LMM:

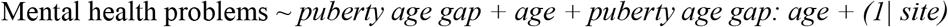

Finally, given the known association of pubertal timing with BMI, SES, and race/ethnicity (Mendle et al., 2019), supplementary analyses repeated primary models while accounting for these variables as covariates (see SI, Appendix S5 and Table S2, for details of these analyses). This approach avoided potential complexity in our primary analyses due to the collinearity of confounding variables with our main variables of interest.

## Results

### Accuracy of puberty age models

All three models were able to provide significant and accurate out-of-sample predictions of age (Table 1, Figure 1). The performance of the hormonal model was better than the physical model in males, while on the contrary, physical puberty age was found to be a better predictor of age than hormonal puberty age in females. Across both sexes, combined puberty age explained the largest degree of variation in chronological age. Non-parametric statistical comparisons showed that the MAE between all models was significantly different (p<0.001). See the correlation between these three models in SI (See Appendix S2 and Figure S1).

**Table 1.** Comparison of the out-of-sample prediction performance of alternative puberty age gap models

|  | Female |  | Male |  |
| --- | --- | --- | --- | --- |
|  | r | mae | r | mae |
| Combined puberty age | 0.58 | 0.65 year | 0.54 | 0.67 year |
| Hormonal puberty age | 0.37 | 0.76 year | 0.45 | 0.72 year |
| Physical development puberty age | 0.55 | 0.67 year | 0.43 | 0.72 year |

r = correlation, mae = mean absolute error

**Figure 1.**
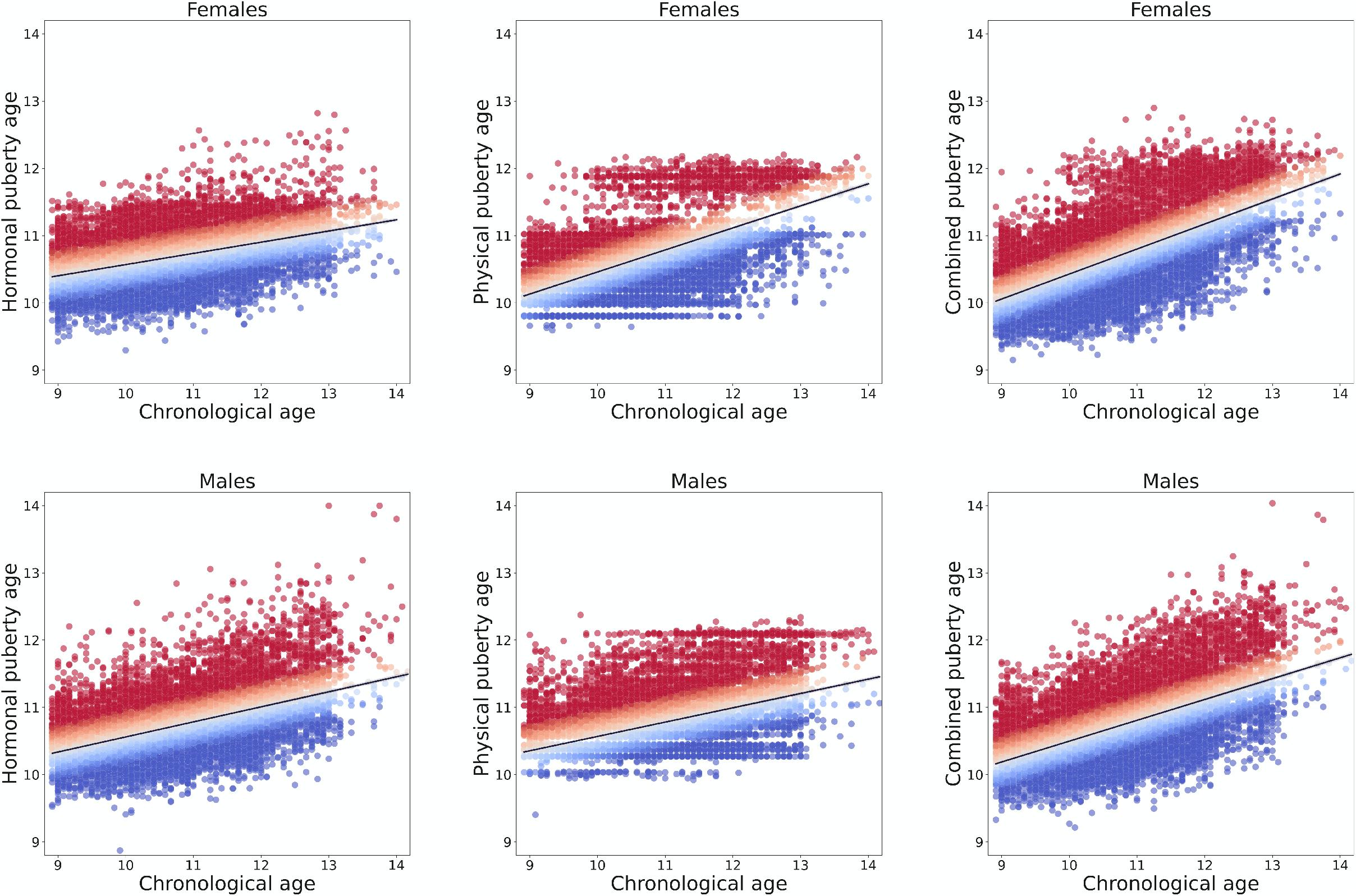
Performance of puberty age gap models. For all three models (combined, physical development and hormonal), the prediction of the age of each individual is plotted against chronological age (in years). Red dots indicate individuals with a positive puberty age gap (early timing) and blue dots indicate individuals with a negative puberty age gap (late timing).

### “Puberty age model” vs. Traditional model

Comparing the out-of-sample prediction performance of the physical puberty age gap with the traditional model of pubertal timing showed that the physical puberty age model was significantly better at modelling the relationship between puberty development and age in previously unseen data (See Table 2). Non-parametric statistical comparisons showed that the MAE between models were significantly different (p<0.001)

**Table 2.** Comparison of the out-of-sample prediction performance of the traditional pubertal timing model and the physical development puberty age model

|  | Female |  | Male |  |
| --- | --- | --- | --- | --- |
|  | *r* | *mae* | *r* | *mae* |
| Traditional pubertal timing | 0.49 | 0.71 | 0.32 | 0.78 |
| Physical development Puberty age Model | 0.55 | 0.68 | 0.43 | 0.72 |

*r* = correlation, *mae* = mean absolute error

#### Pubertal timing measures predicting mental health problems

We investigated the association between each puberty age gap measure (hormonal, physical, and combined) and different dimensions of mental health problems (see Table 3, Figure 2). Hormonal puberty age gap was not significantly associated with any mental health problems in either males or females. Physical puberty age gap, however, was significantly positively associated with all dimensions of mental health problems in males and all dimensions except Anxiety-Depression in females. While similar associations were present for the combined puberty age gap, AIC differences indicated a better model fit for the physical puberty age gap in all cases. Finally, there were no significant interactions between age and any puberty age gap measure in predicting mental health problems in either females or males, suggesting that associations were stable across the age range.

**Table 3.**
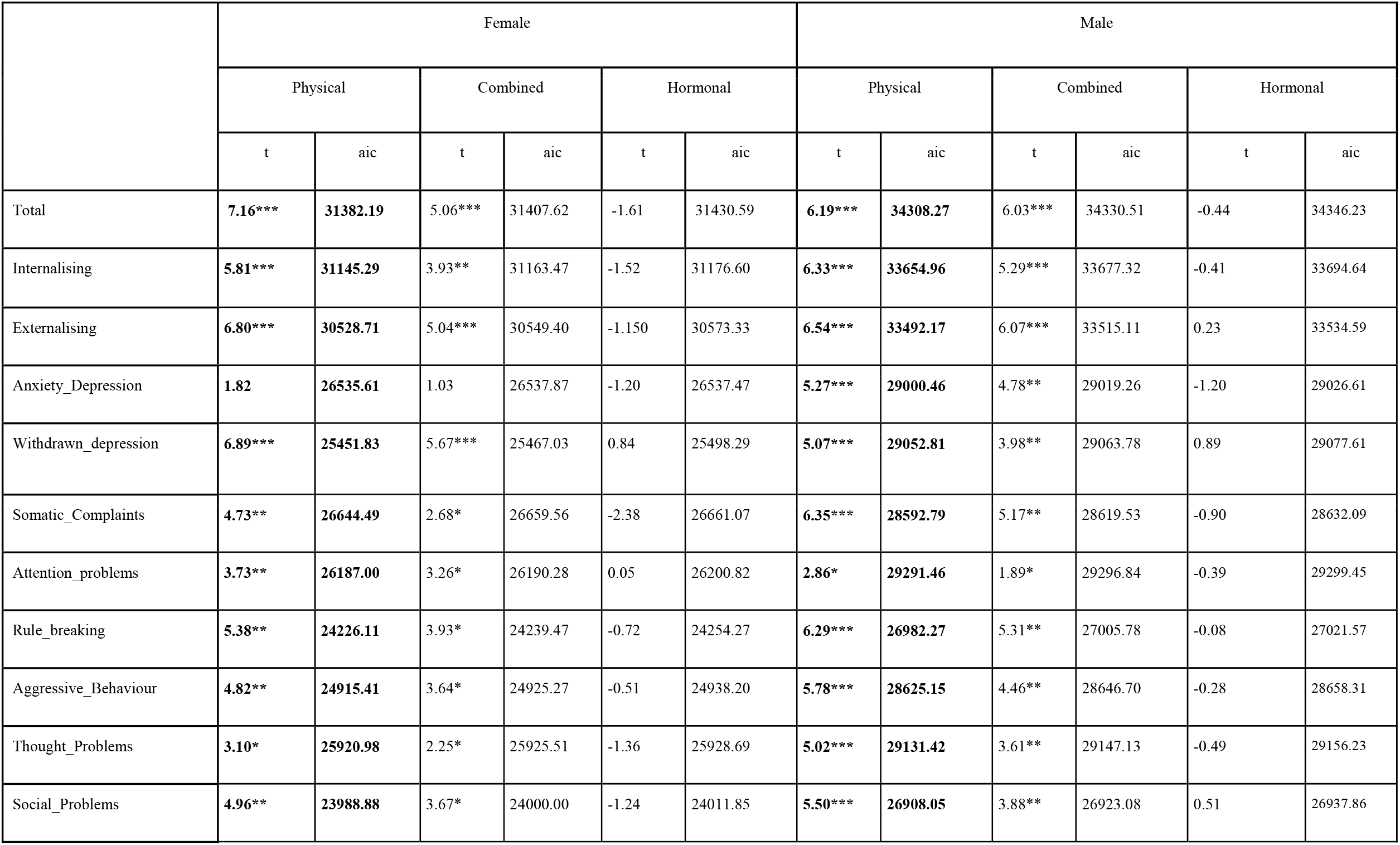
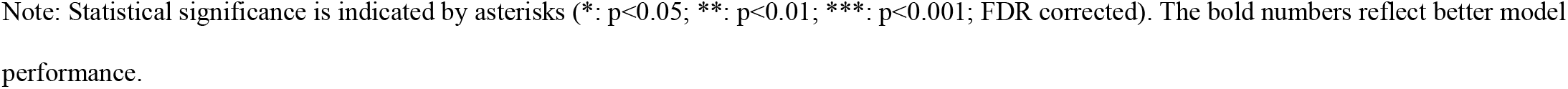
Associations between puberty age gap and mental health problems measured by linear mixed effect regression

|  | Female |  |  |  |  |  | Male |  |  |  |  |  |
| --- | --- | --- | --- | --- | --- | --- | --- | --- | --- | --- | --- | --- |
|  | Physical |  | Combined |  | Hormonal |  | Physical |  | Combined |  | Hormonal |  |
|  | t | aic | t | aic | t | aic | t | aic | t | aic | t | aic |
| Total | **7.16\*\*\*** | **31382.19** | 5.06*** | 31407.62 | -1.61 | 31430.59 | **6.19\*\*\*** | **34308.27** | 6.03*** | 34330.51 | -0.44 | 34346.23 |
| Internalising | **5.81\*\*\*** | **31145.29** | 3.93** | 31163.47 | -1.52 | 31176.60 | **6.33\*\*\*** | **33654.96** | 5.29*** | 33677.32 | -0.41 | 33694.64 |
| Externalising | **6.80\*\*\*** | **30528.71** | 5.04*** | 30549.40 | -1.150 | 30573.33 | **6.54\*\*\*** | **33492.17** | 6.07*** | 33515.11 | 0.23 | 33534.59 |
| Anxiety_Depression | **1.82** | **26535.61** | 1.03 | 26537.87 | -1.20 | 26537.47 | **5.27\*\*\*** | **29000.46** | 4.78** | 29019.26 | -1.20 | 29026.61 |
| Withdrawn_depression | **6.89\*\*\*** | **25451.83** | 5.67*** | 25467.03 | 0.84 | 25498.29 | **5.07\*\*\*** | **29052.81** | 3.98** | 29063.78 | 0.89 | 29077.61 |
| Somatic_Complaints | **4.73\*\*** | **26644.49** | 2.68* | 26659.56 | -2.38 | 26661.07 | **6.35\*\*\*** | **28592.79** | 5.17** | 28619.53 | -0.90 | 28632.09 |
| Attention_problems | **3.73\*\*** | **26187.00** | 3.26* | 26190.28 | 0.05 | 26200.82 | **2.86\*** | **29291.46** | 1.89* | 29296.84 | -0.39 | 29299.45 |
| Rule_breaking | **5.38\*\*** | **24226.11** | 3.93* | 24239.47 | -0.72 | 24254.27 | **6.29\*\*\*** | **26982.27** | 5.31** | 27005.78 | -0.08 | 27021.57 |
| Aggressive_Behaviour | **4.82\*\*** | **24915.41** | 3.64* | 24925.27 | -0.51 | 24938.20 | **5.78\*\*\*** | **28625.15** | 4.46** | 28646.70 | -0.28 | 28658.31 |
| Thought_Problems | **3.10\*** | **25920.98** | 2.25* | 25925.51 | -1.36 | 25928.69 | **5.02\*\*\*** | **29131.42** | 3.61** | 29147.13 | -0.49 | 29156.23 |
| Social_Problems | **4.96\*\*** | **23988.88** | 3.67* | 24000.00 | -1.24 | 24011.85 | **5.50\*\*\*** | **26908.05** | 3.88** | 26923.08 | 0.51 | 26937.86 |

Note: Statistical significance is indicated by asterisks (\*: $p < 0.05$ ; \*\*: $p < 0.01$ ; \*\*\*: $p < 0.001$ ; FDR corrected). The bold numbers reflect better model performance.

**Figure 2.**
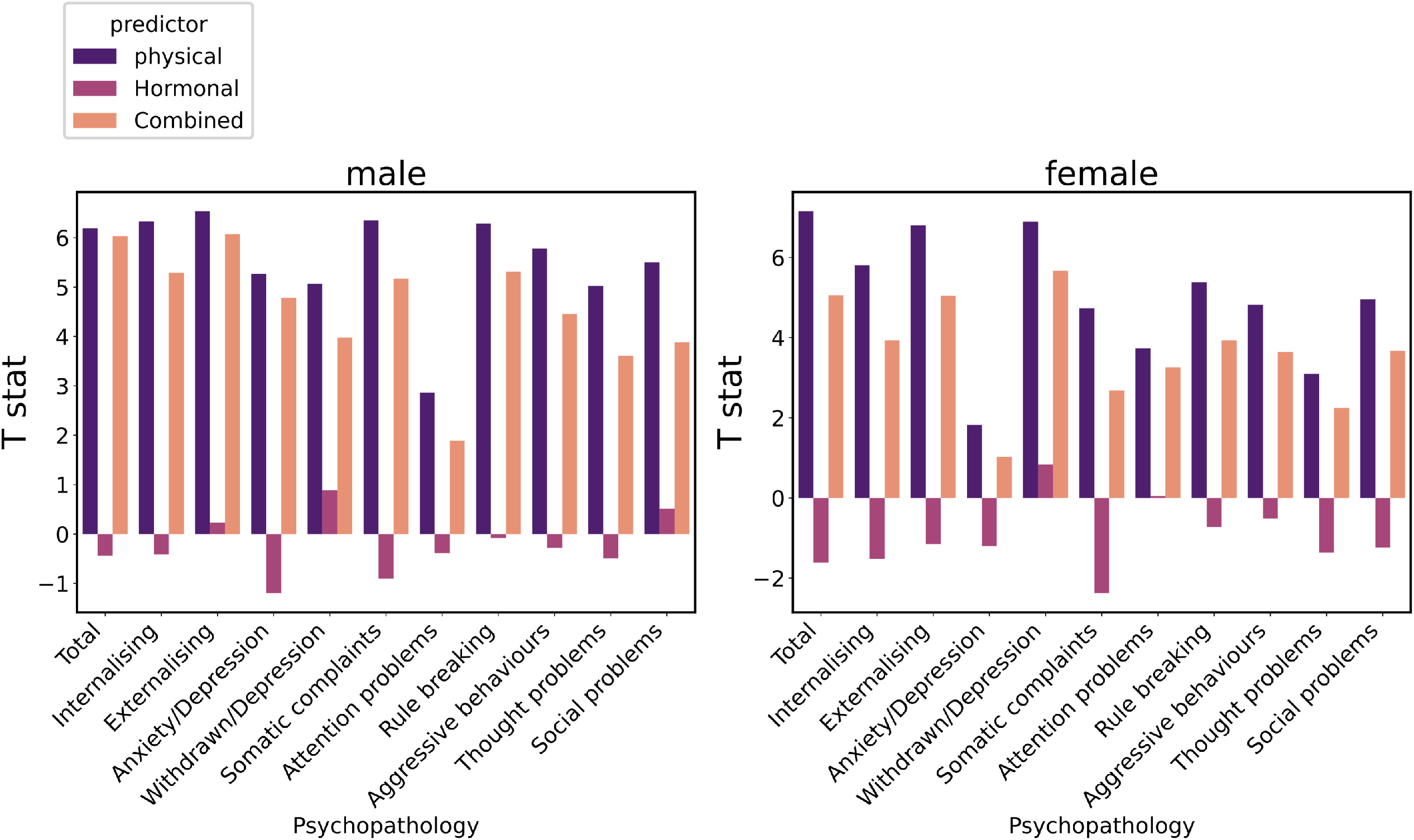
Bar plot showing the T-statistic from the prediction of each dimension of mental health problems from different puberty age models. Hormonal puberty age gap is not significantly associated with any dimension of mental health problems.

## Discussion

In this study, we utilized biological and physical pubertal features and used supervised machine learning to model pubertal timing and its association with mental health problems. Overall, a combined puberty age model predicted age more accurately than the unimodal (i.e., physical, or hormonal features alone). In males, the performance of the hormonal model was better than the physical model, which was not the case for females. This may be due to the fact that the two hormones tested (DHEA and testosterone) are more predominant in males, whilst female-dominant hormones (such as estradiol) were not considered. We showed that our novel pubertal timing model (based on physical pubertal features) had better accuracy than a conventional linear regression-based measurement in both sexes. Findings also highlight that our new method of assessing pubertal timing was significantly associated with most dimensions of mental health problems in males and females. Generally, we found that early pubertal maturation (based on physical and combined physical and hormonal features) was consistently predictive of various dimensions of mental health problems and could hence be an indicative risk factor for general mental health problems.

Findings indicated that our novel normative puberty age models were able to provide significantly accurate out-of-sample predictions of age. This nonlinear modelling approach was found to be more accurate at predicting age than traditional measures of pubertal timing that rely on linear modelling of the relationship between (average) pubertal stage and age. These results highlight the presence of nonlinear relationships between age and pubertal development, consistent with previous literature reporting benefits of other nonlinear methods for calculating pubertal timing(21,31). Additionally, our examination of the collinearity between each of the PDS items showed that, although the items are significantly correlated, the correlations between them are less than 0.35 (for more details, see supplementary section 2). This suggests that each PDS item carried a significant amount of independent information. Our method thus used this independent information in a multivariate design instead of simplifying this information by averaging items to create a single PDS score. Furthermore, our out-of-sample prediction provides an evaluation of the generalizability and replicability of study findings (39). We presented how a model that was trained on 90% of the sample could provide accurate out-of-sample predictions in the remaining 10% of the sample. This provides the opportunity for future research to measure pubertal timing in studies with smaller sample sizes, by taking advantage of this pre-trained normative model. This could potentially alleviate some study biases inherent to small sample sizes.

Beyond its benefits for predicting age, our novel method of assessing pubertal timing was also significantly related to mental health problems in adolescents. Across most models of puberty age, we found that relatively early pubertal timing (i.e., positive puberty age gap) was associated with an increase in most dimensions of mental health problems. This is consistent with the maturation disparity hypothesis, whereby a mismatch between physical development and progression of emotional and cognitive development is purported to increase in those with early pubertal timing, which accounts for difficulties navigating the complexities and challenges of this period and may thus result in greater risk for mental health problems (7, 4). In particular, findings showed that physical puberty age gap out-performed combined and hormonal puberty age gap measures in the prediction of mental health problems in both females and males. Thus, although the prediction accuracy of the combined puberty age model was significantly better than physical puberty age, it was not better at explaining mental health problems. Moreover, we did not find any significant associations between hormonal puberty age gap and mental health problems in males or females. These findings are partially consistent with (40), who reported that pubertal timing measured from hormonal information (testosterone and DHEA) did not predict internalizing symptoms. Consistent with prior studies, these findings may suggest that psychosocial mechanisms have a larger role (in contrast to biological mechanisms indexed by hormones) in predicting mental health problems in early adolescence (14). Likewise, prior work has suggested that earlier physical development impacts social functions such as difficulty maintaining friendship with peers who mature later, and a tendency to associate with older adolescents that engage in more externalizing behaviors (12).

While this study has strengths in its large sample size, use of hormonal assays and physical measurement of puberty, as well as implementation of a novel, generalizable method for assessing pubertal timing, there are limitations that should be addressed in future work. First, the age range of the sample used was rather limited and a wider age range could have captured more variance in pubertal maturation and improved the out-of-sample model prediction of age. Future studies that implement our model in a wider age range encompassing the start to completion of puberty would be able to better capture the complete nonlinear relationship between age and pubertal development. Relatedly, while we did not observe any interactive effects of age with pubertal timing in predicting mental health problems symptoms in early adolescents, it is possible that such effects may be detectable across a larger age range. Further, different measures of puberty age gap may also differentially predict mental health problems in earlier versus later adolescence. Second, this study focused on a limited number of pubertal hormones, and the omission of other hormonal features may have impacted the predictive power of hormonal puberty age gap – particularly in females. Additionally, variations in the time for collecting hormones across the day could have also had an impact on the inferior performance of the hormonal age gap (although these were statistically accounted for). Thus, future studies could incorporate more detailed assessments of hormones to improve model prediction. Although the current study used the parent report version of PDS due to reportedly higher reliability in this particular age range, it would be valuable for future research to replicate this analysis in a sample with a wider age range with self- or clinician-assessed pubertal development.

In conclusion, the current study proposes a smooth nonlinear puberty age model that facilitates generalizable investigations of pubertal timing in future studies. Our findings also highlight the importance of physical pubertal maturation, relative to hormonal changes, for mental health problems during early adolescence. This suggests that psychosocial mechanisms may play an important role in the relationship between early pubertal timing and mental health problems, which has implications for interventions aimed at reducing the risk of the emergence of mental health problems in adolescence.

## Data Availability

This study used data from Adolescent Brain Cognitive Development (ABCD) Study (https://abcdstudy.org), held in the NIMH Data Archive (NDA). This is a longitudinal study which collected data from ~ 11000 children in age 9-10 and follow them through 10 years. A full list of supporters is available at https://abcdstudy.org/federal-partners.html. A listing of participating sites and a complete listing of the study investigators can be found at https://abcdstudy.org/scientists/workgroups/. ABCD consortium investigators designed and implemented the study and/or provided data but did not participate in the analysis or writing of this report. This manuscript reflects the views of the authors and may not reflect the opinions or views of the NIH or ABCD consortium investigators.

## Data Sharing

This study used data from Adolescent Brain Cognitive Development (ABCD) Study (https://abcdstudy.org), held in the NIMH Data Archive (NDA). This is a longitudinal study which collected data from ∼ 11000 children in age 9-10 and follow them through 10 years. A full list of supporters is available at https://abcdstudy.org/federal-partners.html. A listing of participating sites and a complete listing of the study investigators can be found at https://abcdstudy.org/scientists/workgroups/. ABCD consortium investigators designed and implemented the study and/or provided data but did not participate in the analysis or writing of this report. This manuscript reflects the views of the authors and may not reflect the opinions or views of the NIH or ABCD consortium investigators.

## Acknowledgement

Niousha Dehestani was supported by a Deakin University Postgraduate

Research Scholarships (DUPRS). Additionally, this study was supported by the MASSIVE high-performance computing facility (www.massive.org.au).

